# Patterns and Predictors of Dropout in Maternal Continuum of Care: A Comprehensive Analysis in Bangladesh

**DOI:** 10.64898/2026.04.20.26351272

**Authors:** Sabrina Sharmin Priyanka, Md. Sazzad Hossan Sujon, Amita Farzana, Dibbya Prava Dasgupta, Golam Sarower Bhuyan, Nazia Binte Ali

## Abstract

Dropout from essential maternal health services across pregnancy, childbirth, and the postnatal period remains a major barrier to improving maternal and neonatal outcomes in Bangladesh. This study examined stage-specific dropout patterns along the maternal continuum of care and identified factors associated with discontinuation.

We analysed nationally representative data from the Bangladesh Demographic and Health Survey 2022 for 5,162 women with a recent live birth. Dropout from antenatal care, skilled birth attendance, and postnatal care was examined using multivariable logistic regression to estimate adjusted odds ratios and 95% confidence intervals, with comparisons to BDHS 2017–18 and assessment of regional variation.

Only 44% of women received four or more antenatal care visits. Of these, 33% delivered with a skilled birth attendant, and among those receiving both antenatal care and skilled delivery, only 15% received postnatal care within 48 hours. Overall, 57% dropped out before completing adequate antenatal care, with additional dropouts between antenatal care and delivery (10%) and between delivery and postnatal care (18%). Compared with 2017–18, overall dropout from the maternal continuum of care more than doubled in 2022 (5.0% to 11.7%), driven by increased antenatal care dropout, while skilled birth attendance dropout declined and postnatal care dropout increased slightly. Higher maternal education, household wealth, media exposure, and women’s decision-making power were consistently associated with lower odds of dropout, whereas higher birth order increased dropout risk. Substantial regional variation was observed, with the highest overall dropout in Sylhet and the lowest in Khulna.

High dropout from the maternal continuum of care in Bangladesh occurs predominantly at the antenatal care stage and is shaped by socioeconomic status, birth order, women’s access to information, and regional disparities. Strengthening early antenatal engagement and women’s decision-making autonomy is critical to improving continuity of maternal care and reducing preventable maternal and neonatal risks.

## Introduction

Despite global efforts, most global maternal and neonatal deaths occur in low and middle-income countries (LMICs), and the majority of these countries are not on track to meet the Sustainable Development Goals (SDG) targets by 2030. In LMICs alone, the maternal mortality ratio stood at 430 deaths per 100,000 live births in 2020 [1] and 2.3 million children died within their first month of life in 2022, underscoring the urgent need to improve maternal and neonatal care in these regions [2, 3]. The Continuum of Care (CoC) for maternal, newborn, and child health (MNCH) includes an integrated service delivery model starting from antenatal care (ANC) during pregnancy, skilled birth attendance during childbirth, and postnatal care (PNC) in the postpartum period [4–6]. This model aims to improve maternal and newborn care, recognizing the interdependence of maternal and newborn health and highlighting the need for continuation of care during the vulnerable period of pregnancy, childbirth, and the immediate postnatal period [6]. ANC is the care provided by skilled health-care professionals to pregnant women in order to ensure the best health conditions for both mother and baby during pregnancy [7]. ANC visits can detect, address, and prevent complications during pregnancy, enhancing the likelihood of receiving appropriate care during childbirth [7, 8]. Skilled attendance during labor and delivery is crucial for a safe birth and essential newborn care, significantly reducing the risk of mortality for both mother and baby from delivery-related complications [9]. On the other hand, postnatal care (PNC), immediately after birth and continuing for up to six weeks, is essential to prevent maternal death from excessive bleeding, infections, and hypertensive disorder of pregnancy as well as prevent neonatal death from major causes such as complications of prematurity, birth asphyxia, and sepsis [10, 11]. Therefore, successful completion of the MNCH CoC can not only lower maternal and neonatal mortality and morbidities from preventable causes but also reduce long-term physical and psychological complications for the mothers and babies, thereby reducing disabilities, long-term health care costs and facilitating progress towards SDG goals [12].

The high dropout rate for MNCH CoC is a global public health challenge. Specifically, LMICs observed the highest dropout rates for MNCH CoC, moving sequentially from ANC to institutional delivery and from institutional delivery to PNC services [6, 12, 13]. In Africa, for example, the rates range from 8% in Ghana to 31% in Nigeria [14, 15]. In India, due to substantial regional variation, completion rates range from 12% to 81%, with the highest number of dropouts in the CoC observed at ANC, with a loss of nearly 38% [4]. In Nepal, more than half of the pregnant women in the country do not complete the CoC [16]. The dropout patterns in Africa and Southeast Asia suggest that the transition from ANC to delivery care (including skilled birth attendance) is a critical point where many women fall out of the CoC, emphasizing the need for strategic interventions [17]. Among the socio-economic factors, poverty, distance to health facilities, limited information, poor service quality, and prevailing cultural beliefs and practices have been identified as the major causes of overall maternal CoC dropouts in LMICs [13, 17]. Studies also identified maternal characteristics such as age at first birth, the number of children, mother’s educational attainment, household wealth, place of residence, women’s autonomy, and exposure to mass media, being associated with CoC. In addition, this study examined regional variations in CoC dropout to capture geographic patterns of maternal healthcare utilization across diverse settings, thereby providing evidence to inform policy decisions and the development of targeted interventions. Understanding these geographic variations is crucial, as disparities in healthcare infrastructure, service availability, and sociocultural practices across regions can significantly influence women’s ability and willingness to complete the continuum of maternal care.

Bangladesh is one of the LMICs that has made significant progress in reducing maternal and child mortality over the last few decades: 322 per 100 000 live births in 2001 to 194 per 100 000 live births in 2010 [18–20]. However, the country has observed a lag in the decline of maternal mortality since then, as depicted by 196 maternal deaths per 100,000 in 2016 [21]. One of the major reasons for this lag in decline is the high dropout from MNCH CoC. In Bangladesh, 41% of women used four and above ANC, 65% had institutional delivery, and 61% of women had PNC visits [22]. The completion rate of the CoC in Bangladesh is low, with only 18% of women completing all seven MNCH care interventions during their reproductive lifespan [23]. The high dropout rate from the maternal CoC in Bangladesh is a significant public health concern with far-reaching consequences such as missed opportunities to identify and timely manage preventable causes of maternal and newborn mortality and morbidities [23, 24], exacerbating existing health inequities in maternal and newborn deaths within the country [23, 25, 26] and poor maternal and newborn health outcomes, leading to increased healthcare costs, loss of productivity, and long-term economic burdens on families and the healthcare system [23, 27].

This study assessed patterns and factors associated with maternal CoC dropout in Bangladesh. The primary objective was to investigate the extent of dropout within and across stages of care, from pregnancy to delivery and the postnatal period for mothers. The secondary objective was identifying factors associated with dropouts at different continuum stages. Findings from this study inform policies and design targeted interventions to improve maternal CoC in Bangladesh and globally.

## Materials and Methods

### Study design

We did a secondary analysis of the Bangladesh Demographic and Health Survey (BDHS) 2022 data [28]. The national survey aimed to provide reliable demographic and health estimates for Bangladesh, covering urban and rural areas of eight administrative divisions. The BDHS 2022s sampling design and questionnaires can be found in the final report [28]. Briefly, the survey employed a two-stage stratified sample design. The primary sampling unit (PSU) was an enumeration area (EA) with an average of 120 households. A total of 675 EAs were selected, including 237 urban and 438 rural areas from the 2011 Population and Housing Census [29]. In the second stage, a systematic sample of 30 households were selected from each EA. For this analysis, we included 5,162 mothers with live births in the past three years of the survey. In cases of multiple births, we used data from the youngest child. In addition, we performed a descriptive comparison of maternal CoC dropout trends using data from both BDHS 2017–18 and BDHS 2022, to assess changes over time across key care stages (ANC, SBA, and PNC).

### Study Variables

The outcome variable of the study is a dropout from the maternal CoC. This included four or more antenatal care, delivery with a skilled birth attendance (defined as delivered by either qualified a doctor, nurse, midwife, family welfare visitor (FWV), community skilled birth attendant, sub assistant community medical office or community health care provider), and postnatal care (PNC) within 48 hours for mothers (either in a facility or at home), examining at which stages mothers might have dropped out of the recommended care continuum.

This study used the definition of variables regarding dropout from the maternal care continuum from previous studies [12, 13]. ANC dropout was defined as a woman who had less than four ANC visits during her most recent childbirth. Skilled birth attendant (SBA) dropouts included women who had four or more ANC visits but did not seek an SBA during delivery, meaning the delivery was not conducted by healthcare professionals such as midwives, nurses, doctors, and/or health officers. A PNC dropout was a woman who had four or more ANC visits and had SBA during delivery but did not receive PNC visits within the first 48 hours of delivery. Dropouts from maternal CoC included women who dropped out of ANC, SBA, and PNC. It included dropouts from the entire continuum of maternal care, from pregnancy to post-delivery.

Our exposure variables included sociodemographic factors, reproductive and obstetric factors, and women’s empowerment. Sociodemographic factors included the mother’s age (<20, 20-34, 35-49), religion (Muslim and others), level of education (no education, primary, secondary and higher), division (Sylhet, Barisal, Chittagong, Dhaka, Khulna, Mymensingh and Rajshashi), household income (poorest, poorer, middle, richer, richest), area of residence (rural and urban), mobile phone ownership (yes or no), and media exposure (yes or no). Reproductive and obstetrics factors, included birth order (one birth, two births and 3 or more births), and the mother’s body mass index (BMI) (underweight-<18.5 kg, normal −18.5 kg to 24.9 kg, overweight- 25 to 30kg, obesity- <30 kg). Women’s empowerment was defined as mothers participating in all three specified household decision making regarding their health care, household purchases, and/or visits to family or relatives.

### Statistical analysis

The analysis was performed using STATA software, version 17 and R-Studio. First, we examined the socio-demographic characteristics of the study participants. Descriptive statistics were used to compare dropout rates between BDHS 2017–18 and BDHS 2022. Then, we calculated the rates of dropouts in each stage of CoC. Then we estimated adjusted odd ratios (aORs) and 95% confidence intervals (CIs) for the associations between dropout from each stages of the continuum of care (CoC) and mother’s age, religion, level of education, division, household income, area of residence, mobile phone ownership, media exposure, birth order, mother’s body mass index (BMI) and household decision-making power using four sequential multivariable generalized linear models (GLM) with binomial distribution and logit link. Each model targeted a specific stage of CoC dropout: Model I for ANC dropout, Model II for SBA dropout, Model III for PNC dropout, and Model IV for dropout from the overall maternal CoC. Each dropout was coded as ‘1’ for dropout and ‘0’ otherwise. We did not include survey weights in these models.

The models were of the form

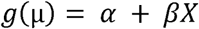

In examining dropout patterns in the maternal Continuum of Care (CoC) using a Generalized Linear Model (GLM) with a logit link function, we model the log odds of dropping out at various stages such as ANC to delivery or delivery to PNC. The link function, 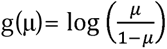 transforms the probability μ of dropout into log odds. The model’s intercept, α, represents the baseline log odds of dropout when all predictor variables are zero, setting a fundamental reference point.

Each coefficient β in the model describes how the corresponding predictor X factors influences the log odds of dropping out of the maternal CoC. The predictor variables X, like maternal education, household wealth, age at first birth, media exposure, and cultural influences affect the log odds of CoC dropout. Positive coefficients indicate an increased risk of dropout with the respective predictors. These predictors are chosen based on their demonstrated or suspected impact on healthcare continuation and are controlled for other variables to isolate their effects.

Finally, we explored regional variations in adjusted odds ratios (AORs) for maternal healthcare dropout (ANC, SBA, PNC and full continuum of care) among the administrative divisions of Bangladesh using spatial analysis.

### Ethical consideration

The BDHS 2022 was carried out by National Institute of Population Research and Training (NIPORT), affiliated with Mitra and Associates, ICF, and icddr,b. During the survey, the implementing organizations took informed written consent from the study participants. We did a secondary analysis of the publicly available de-identified BDHS 2022 dataset, which is not human-subject research.

## Results

### Demographic characteristics of respondents

The survey involved 5,162 participants, predominantly aged 20-34 years (89.2%). More than half of the participants completed secondary education (Table 1). Around two-thirds of the participants owned mobile phones (69.9%), and 56.6% had media exposure. 67% lived in urban areas. Over half (56.6%) had media exposure. The majority of the participants were Muslim (91.4%).

**Table 1.** Demographic characteristics of the respondents by dropout from the continuum of care (CoC).

|  |  | Dropout from continuum of care (CoC) |  |  |
| --- | --- | --- | --- | --- |
| Variable | Total<br>N=5162 | No<br>N (%) | Yes<br>N (%) | P-Value |
| Age |  |  |  |  |
| >20 | 1150 | 1026 (89.2%) | 124 (10.8%) | <0.001 |
| 20-34 | 3748 | 3322 (88.6%) | 426 (11.4%) |  |
| 35+ | 264 | 210 (79.6%) | 54 (20.5%) |  |
| Religion |  |  |  |  |
| Islam | 4720 | 4150 (87.9%) | 570 (12.1%) | 0.006 |
| Others | 442 | 408 (92.3%) | 34 (7.7%) |  |
| Women’s Education |  |  |  |  |
| No education | 273 | 199 (72.9%) | 74 (27.1%) | <0.001 |
| Primary | 1179 | 957 (81.2%) | 222 (18.8%) |  |
| Secondary | 2729 | 2453 (89.9%) | 276 (10.1%) |  |
| Higher education | 981 | 949 (96.7%) | 32 (3.3%) |  |
| Partners' education |  |  |  |  |
| No | 768 | 596 (77.6%) | 172 (22.4%) | <0.001 |
| Yes | 4394 | 3962 (90.2%) | 432 (9.8%) |  |
| Division |  |  |  |  |
| Barishal | 554 | 489 (88.3%) | 65 (11.7%) | <0.001 |
| Chattogram | 893 | 815 (91.3%) | 78 (8.7%) |  |
| Dhaka | 767 | 681 (88.8%) | 86 (11.2%) |  |
| Khulna | 587 | 560 (95.4%) | 27 (4.6%) |  |
| Mymensingh | 631 | 527 (83.5%) | 104 (16.5%) |  |
| Rajshahi | 523 | 468 (89.5%) | 55 (10.5%) |  |
| Rangpur | 599 | 519 (86.6%) | 80 (13.4%) |  |
| Sylhet | 608 | 499 (82.1%) | 109 (17.9%) |  |
| Wealth |  |  |  |  |
| Poorest | 1052 | 818 (77.8%) | 234 (22.2%) | <0.001 |
| Poorer | 1045 | 895 (85.7%) | 150 (14.4%) |  |
| Middle | 1034 | 916 (88.6%) | 118 (11.4%) |  |
| Richer | 1023 | 949 (92.8%) | 74 (7.2%) |  |
| Richest | 1008 | 980 (97.2%) | 28 (2.8%) |  |
| Birth order |  |  |  |  |
| 1st Birth | 1931 | 1789 (92.7%) | 142 (7.4%) | <0.001 |
| 2 Births | 1812 | 1620 (89.4%) | 192 (10.6%) |  |
| 3 or more births | 1419 | 1149 (81%) | 270 (19%) |  |
| Residence |  |  |  |  |
| Rural | 1705 | 1580 (92.7%) | 125 (7.3%) | <0.001 |
| Urban | 3457 | 2978 (86.1%) | 479 (13.9%) |  |

| BMI |  |  |  |  |
| --- | --- | --- | --- | --- |
| Underweight (<18.5) | 382 | 315 (82.5%) | 67 (17.5%) | <0.001 |
| Normal (18.5- 24.9) | 1483 | 1299 (87.6%) | 184 (12.4%) |  |
| Overweight (25-30) | 585 | 541 (92.5%) | 44 (7.5%) |  |
| Obesity (<30) | 130 | 120 (92.3%) | 10 (7.7%) |  |
| Missing | 2582 | 2283 (88.4%) | 299 (11.6%) |  |
| Have Mobile |  |  |  |  |
| No | 1553 | 1262 (81.3%) | 291 (18.7%) | <0.001 |
| Yes | 3609 | 3296 (91.3%) | 313 (8.7%) |  |
| Decision making power in the household |  |  |  |  |
| No | 2334 | 2041 (87.5%) | 293 (12.6%) | 0.083 |
| Yes | 2828 | 2517 (89%) | 311 (11%) |  |
| Media exposure |  |  |  |  |
| No | 2240 | 1867(83.4%) | 373(16.7%) | <0.001 |
| Yes | 2922 | 2691(92.1%) | 231(7.9%) |  |

We observed higher dropout rates from the maternal CoC among women aged 35 years and above (20.5%) and among women without any formal education (27.1%). Regionally, the Sylhet division had the highest dropout rate at 17.9%, while Mymensingh had the lowest at 4.6%. A notable disparity was observed in dropout rates between the poorest and richest wealth quintiles (22.2% vs. 2.8%). Higher dropouts were also seen in children of higher birth order compared to first-borns. Urban women had a higher dropout rate (13.9%) compared to non-Muslim mothers (7.3%). CoC dropouts were less frequent among women who owned mobile phones (8.7%) and those with media exposure (7.9%).

### The proportion of dropouts from the continuum of maternity care

The flowchart illustrates the stepwise dropout rates at each stage of maternal CoC among 5,162 participants (Fig 1). It indicates that 44% of women received four or more ANC visits. Of those, 33% had SBA during delivery. Among the women who received both four or more ANC visits and SBA, only 15% received PNC. This reflects a 57% dropout rate before completing four ANC visits, a further 10% dropout between ANC and SBA, and an additional 18% dropout from delivery to PNC.

**Fig 1.**
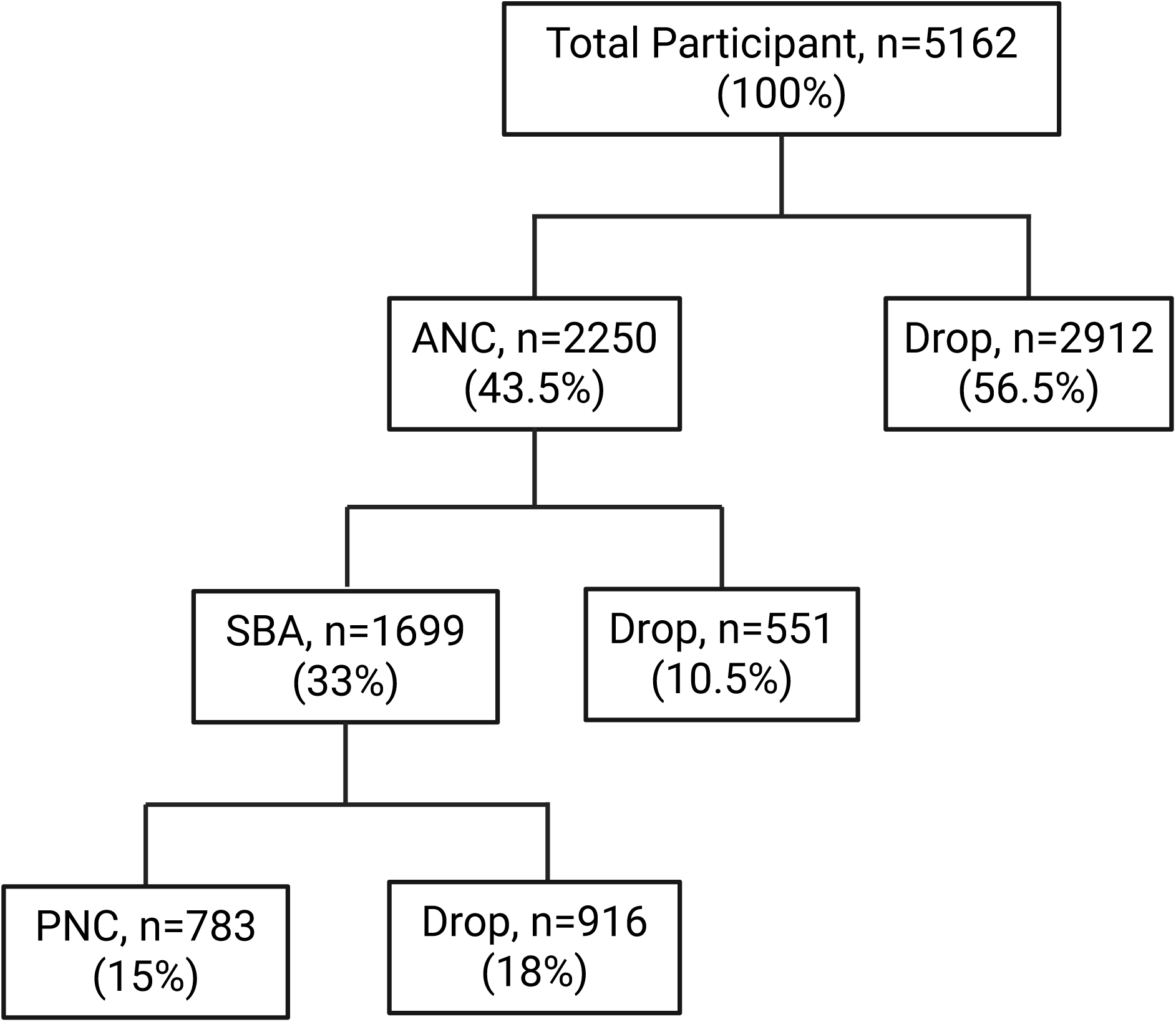
Flowchart of the continuum of care across the maternal and newborn healthcare of women for the most recent birth, Bangladesh, BDHS, 2022. Flow diagram illustrating progression through the maternal continuum of care for the most recent birth in BDHS 2022 (n = 5,162). The chart presents the number and proportion of women receiving four or more antenatal care visits (ANC), skilled birth attendance (SBA), and postnatal care within 48 hours (PNC), along with stage-specific dropout at each transition point.

We compared dropout rates of CoC between BDHS 2017–18 and BDHS 2022 (Fig 2). Overall dropout from the CoC doubled from 5.0% to 11.7% between 2017–18 and 2022. ANC dropout increased from 50.5% to 56.5% during this period. In contrast, there was a decline in the SBA delivery dropout from 16.5% to 10.5% during this period. We also observed increased PNC dropouts from 16.6% to 18% from 2017–18 to 2022.

**Fig 2.**
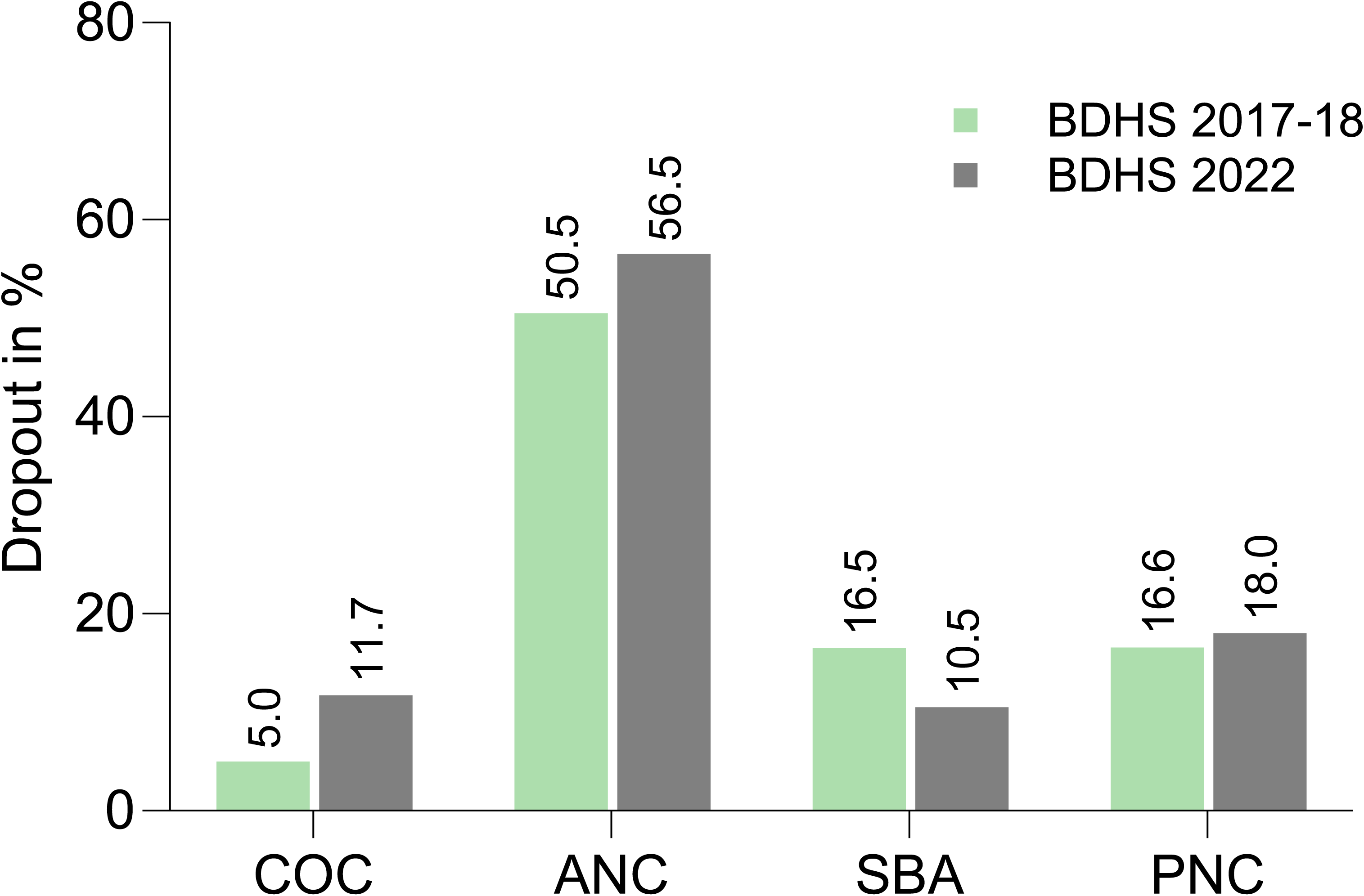
Trends in dropout from maternal continuum of care (CoC): A Comparison of BDHS 2017–18 and 2022. Comparison of stage-specific and overall dropout rates across the maternal continuum of care between BDHS 2017–18 and BDHS 2022. Percentages represent dropout at the ANC, SBA, and PNC stages, as well as cumulative dropout from the full continuum, demonstrating temporal changes in service discontinuation.

### Factors associated with four or more ANC dropouts

We estimated multivariable adjusted associations between sociodemographic factors, reproductive and obstetric characteristics, and women’s decision-making power with dropout from four or more ANC (Table 2). Our multivariable adjusted analysis showed that women with higher education levels had lower odds of ANC dropout compared to women with no formal education (aOR: 0.61; 95% CI: 0.38-0.98). Wealth quintiles were negatively associated with ANC dropout; women in the richer quintiles were less likely to dropout rates from ANC compared to the poorest quintile; aOR for the richer quintile was 0.68 (95% CI: 0.5-0.92) and for the richest quintile was 0.45 (95% CI: 0.32-0.63). Ownership of mobile phones and media exposure during pregnancy reduced the odds of ANC dropout by 33%. In contrast, higher birth order was associated with increased ANC dropout (aOR: 1.4; 95% CI: 1.1–1.77). Women who make their own healthcare decisions had lower odds of dropout from ANC (aOR: 0.82; 95% CI: 0.69-0.97).

**Table 2:**
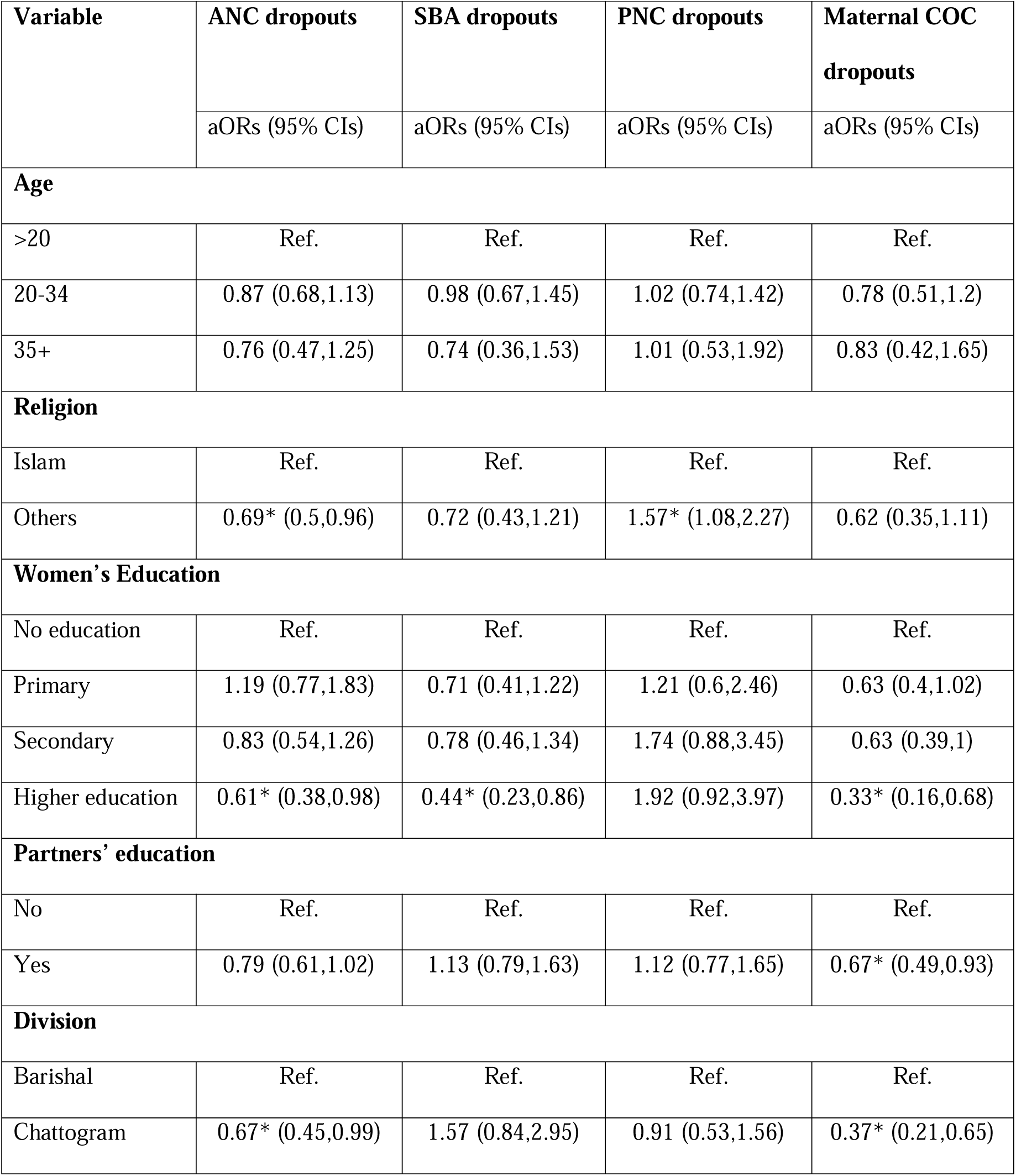
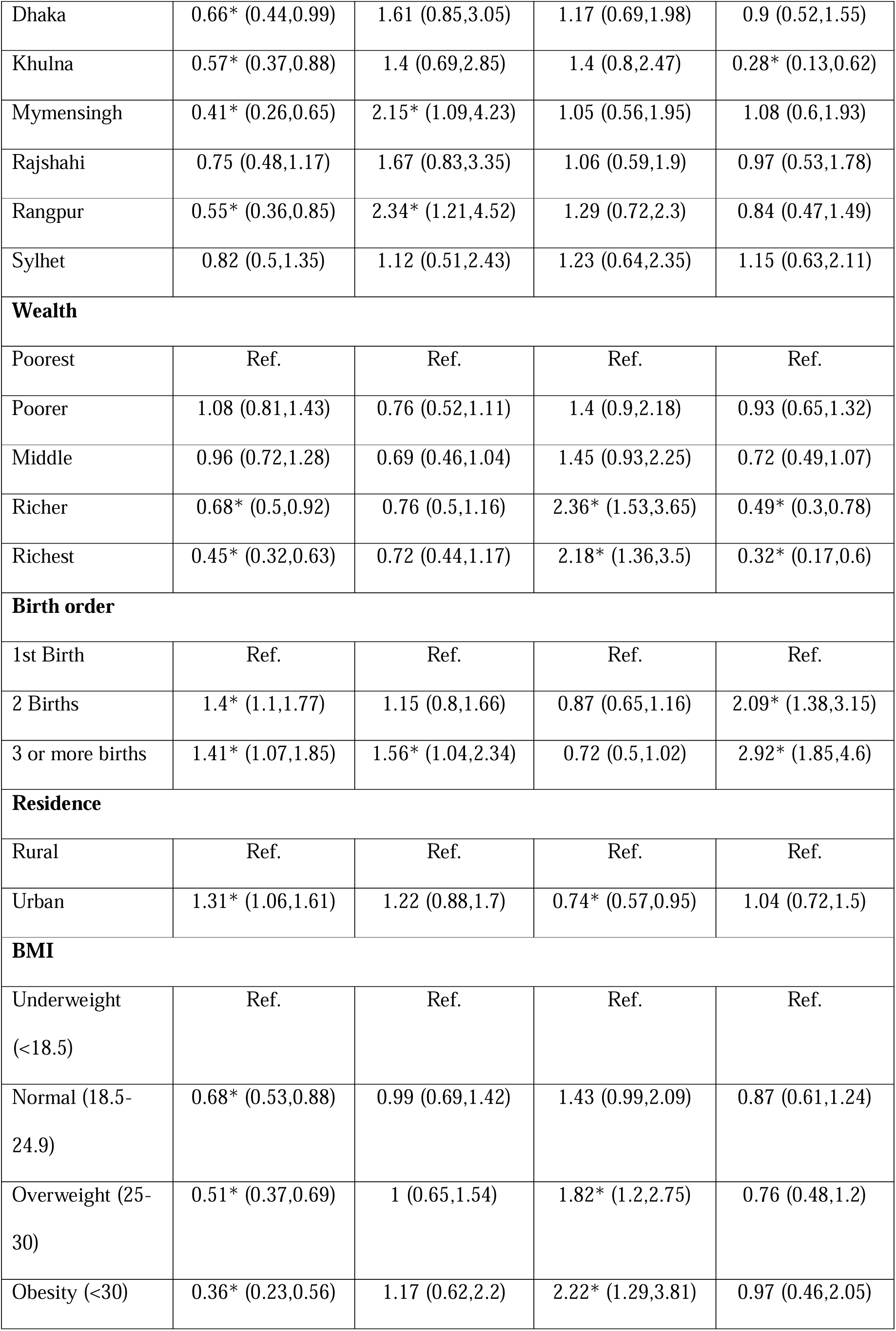
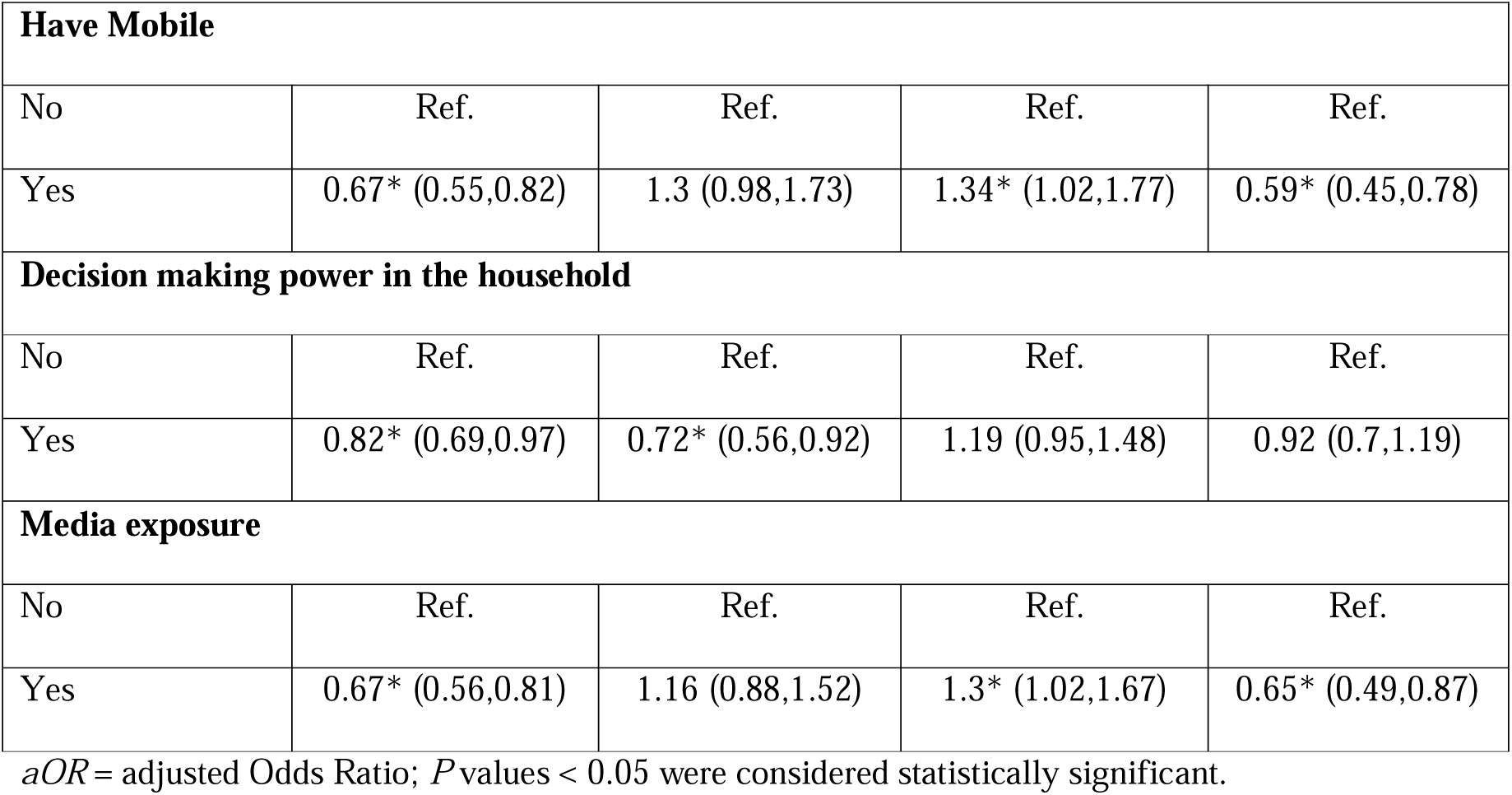
Factors associated with ANC, SBA, PNC and dropout from the maternity care continuum.

### Factors associated with the SBA dropout

Then we explored multivariable adjusted associations between sociodemographic, reproductive, and women’s decision-making factors with dropout from SBA during delivery among women who received four or more ANC (Table 2). We found that women having higher education were less likely to drop out of SBA delivery compared to women without any formal education conditional on other sociodemographic, reproductive, and decision-making factors (aOR: 0.44; 95% CI 0.23-0.86). We also found that women’s decision-making power reduced odd of SBA dropout by 72%. In contrast, having a higher birth order increased the odds of dropout from SBA.

### Factors associated with PNC dropout

We then examined the multivariable adjusted association between sociodemographic, reproductive, and women’s decision-making factors and PNC dropouts among women who have received care four or more ANC and SBA during delivery (Table 2). Surprisingly, we found that women from wealthier households had higher odds of PNC dropout compared to women from the poorest quintile. Additionally, women from the urban areas had a lower odd of PNC dropout compared to their rural counterparts (aOR: 0.74; 95% CI 0.57-0.95).

### Factors associated with dropout from the maternal CoC

Model IV presents multivariable adjusted associations between sociodemographic, reproductive and women’s decision-making factors with dropout from CoC (ANC, SBA during delivery, and PNC). We found women who completed higher education had lower odds of CoC dropout compared to women without any formal education (aOR: 0.33; 95% CI: 0.16-0.68). Women from richer (0.49, 0.3-0.78) and the richest (0.32, 0.17-0.6) wealth quintiles had lower odds of dropout compared to those from the poorest quintile. Owning a mobile phone (aOR:0.59, 95%CI: 0.45-0.78) and having media exposure (0.65, 0.49-0.87) significantly reduced the odds of dropout from CoC. Additionally, women having their second birth (aOR: 2.09, 95% CI: 1.38-3.15), third birth (aOR: 2.92, 95%CI: 1.85-4.6), or more show had higher odds of drop out from CoC compared to those having their first birth.

In Fig 3, we adjusted all model outputs to ensure the average of each model matched the overall average. This makes it easier to compare across divisions while keeping the differences between models intact. The heatmap reveals substantial variation in dropout rates, with the highest ANC dropouts observed in Sylhet (20%) and Chattogram (18%), indicating significant gaps in early maternal engagement. SBA dropouts were most pronounced in Mymensingh (≈19%) and Rangpur (17%), suggesting barriers to skilled delivery services in these regions. PNC dropouts were concentrated in Dhaka and Mymensingh (15%–18%), while the overall CoC dropout was highest in Sylhet (19%) and lowest in Khulna (7%). Overall, divisions such as Khulna and Barishal demonstrated better service retention across the continuum, whereas Sylhet, Mymensingh, and Chattogram consistently experienced higher dropouts, highlighting persistent regional inequities and the need for targeted interventions to improve continuity of maternal care.

**Fig 3.**
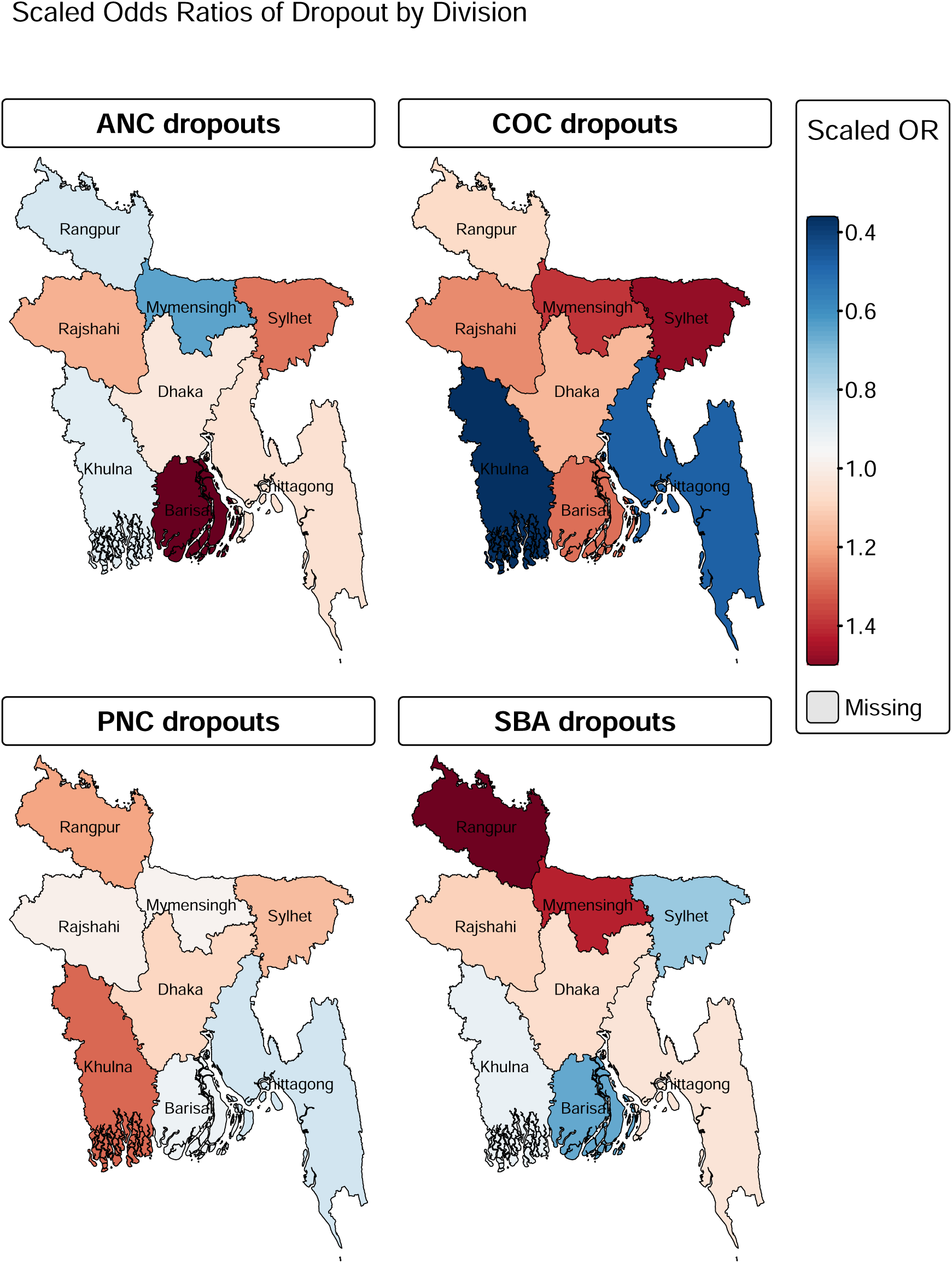
Percentage of dropout in all stages of care by region using heatmap. Heatmap showing regional variation in the percentage of dropout at each stage of the maternal continuum of care (ANC, SBA, PNC, and overall CoC) across administrative divisions of Bangladesh based on BDHS 2022 data. Color intensity reflects the relative magnitude of dropout by region.

## Discussion

Our study provides essential insights into the patterns and determinants of dropout from maternal CoC in Bangladesh. We found significant dropouts across all stages of CoC, with a high dropout from four ANC visits. Among sociodemographic factors explored in this study, we found that women with higher educational attainment, higher household wealth, access to a mobile phone, and media exposure were less likely to drop out from the CoC compared to their counterparts. In contrast, women with higher birth orders were more likely to drop out from CoC compared to women having their first birth. There were regional variations in the dropout from CoC, with Sylhet and Rajshahi divisions having higher ANC dropouts, and Mymensingh and Rangpur divisions having higher SBA dropouts compared to the Barishal division.

Our results showed that more than half of the women did not receive four ANC visits during their last birth, which is the first and most critical point of engagement with the health system. Although coverage of ANC by medically trained provider increased from 82% in 2017-18 to 88% in 2022 in Bangladesh, only 41% of pregnant women received four or more visits in 2022 and this number declined by 5% from 2017-18 [8, 22, 28]. Studies from Pakistan and Tanzania found that four or more ANC visits were associated with increased continuation to SBA and PNC, highlighting the potential negative implications of ANC dropout on subsequent care-seeking and maternal CoC [5, 30–32]. Evidence further indicates that poor quality of ANC in LMICs, including shortages of skilled providers, limited diagnostic capacity, inadequate counseling, poor communication, and lack of respectful care [7], may discourage women and their families from facility delivery and PNC[25, 33]. Future research should explore innovative strategies to not only improve the quality of ANC but also leverage its potential as a platform to engage mothers and encourage continued care-seeking for delivery and PNC services.

We also found that women’s higher educational attainment, access to a mobile phone, and exposure to media were negatively associated with maternal CoC dropout. Evidence suggests that women’s education and access to mobile phones were associated with greater decision-making power and higher utilization of maternal health services, including ANC and SBA [13, 25, 34]. Previous studies from Bangladesh [25], Nepal [16], and Pakistan [35] have similarly found that media exposure correlates with enhanced women’s decision-making autonomy and reduced maternal CoC dropout. These findings underscore the potential of mobile technology to strengthen maternal CoC by delivering health messages, promoting awareness of CoC benefits through digital campaigns, providing SMS reminders for ANC and PNC visits, offering telemedicine services for ANC and PNC in remote areas, and establishing support networks for pregnant women and new mothers[31, 36, 37]. As mobile phone coverage continues to grow in Bangladesh and other LMICs, this platform offers substantial promise for improving access to maternal health services, an area that warrants further research[38].

Our study found significant geographical and socio-economic disparities in maternal CoC dropouts in Bangladesh. Notably, Sylhet and Barisal had high ANC dropout rates, while Mymensingh and Rangpur showed higher dropout from SBA. We also found that women in rural areas were generally more likely to discontinue CoC compared to urban women. Interestingly, ANC dropouts were higher in urban areas compared to rural areas. These findings indicate the presence of context-specific barriers affecting access to ANC, SBA, and PNC services, which require targeted interventions. For example, studies in urban slums of Bangladesh have identified poor service quality as one of the reasons for lower ANC care seeking [39]. In contrast, in hard-to-reach regions of Sylhet and Barisal, distance to facilities, lack of transportation, and cultural or religious beliefs in conservative communities appear to be major contributors to the lack of access to maternal health services, which need further exploration[39, 40]. In the absence of provision of health insurance, poverty and high out-of-pocket costs of care seeking may further hinder maternal care seeking[12, 23, 25]. Given this context, future research should explore interventions tailored to specific local needs, such as mobile clinics, outreach programs, and transportation stipends to improve geographic accessibility in remote and rural areas [18, 41]. To address financial barriers to care seeking, interventions such as community-based health insurance and conditional cash transfers can be examined in future studies [42, 43]. Finally, engaging community leaders, including religious figures, schoolteachers, and health volunteers, may help overcome sociocultural barriers that limit women’s access to maternal health services and should be evaluated in future studies[14, 18].

Our study has several strengths and limitations. Key strengths of our study include the use of a national survey dataset, which provides comprehensive insights into maternal CoC dropouts across Bangladesh. However, this study also has several limitations. Notably, we were unable to assess the quality of ANC, SBA, and PNC services, which may be significant contributors to CoC dropout. Additionally, we did not examine the social, cultural, and financial barriers to maternal care-seeking behaviors. All these factors require further exploration in future research.

In conclusion, our study identified high dropouts from the maternal CoC in Bangladesh, with the majority of the dropouts happening at the first point of contact of ANC. Maternal education, access to mobile phones and media, and higher household wealth were associated with lower CoC dropouts. In contrast, dropout rates were higher in rural areas, and we observed notable geographic disparities across administrative divisions. Given the critical role of ANC in shaping subsequent care-seeking behaviors and the promise of mHealth interventions, both should be explored as platforms for generating demand and improving continued engagement with maternal health services. Additionally, future research should explore contextual barriers at each stage of ANC, SBA, and PNC, including issues related to service quality, geographic accessibility, and socio-cultural and economic factors, and develop and evaluate targeted interventions to address these challenges.

## Data Availability

All data produced in the present work are contained in the manuscript

